# Detection of intratumoral hypoxia in primary breast cancer using photoacoustic imaging

**DOI:** 10.64898/2026.08.20.26360033

**Authors:** Hanako Shimizu, Masahiro Kawashima, Masako Kataoka, Aya Yoshikawa, Yasufumi Asao, Yasuhide Takeuchi, Masahiro Takada, Susumu Saito, Masakazu Toi, Norikazu Masuda

**Affiliations:** Department of Breast Surgery, Graduate School of Medicine, Kyoto University, Kyoto, Japan; Preemptive Medicine and Lifestyle-related Disease Research Center, Kyoto University Hospital, Kyoto, Japan; Luxonus Inc., Kawasaki, Japan; Department of Diagnostic Pathology, Graduate School of Medicine, Kyoto University, Kyoto, Japan; Department of Breast Surgery, Kansai Medical University, Hirakata, Japan; Department of Plastic and Reconstructive Surgery, Graduate School of Medicine, Kyoto University, Kyoto, Japan; Tokyo Metropolitan Cancer and Infectious Disease Center, Tokyo, Japan; Center for Cancer Immunotherapy and Immunobiology, Kyoto University, Kyoto, Japan

## Abstract

**Background:** Tumor hypoxia and abnormal vasculature are closely associated with aggressiveness in solid tumors. Therefore, noninvasive assessment of these features in primary breast cancer is needed. Photoacoustic (PA) imaging is an emerging modality that enables real-time visualization of vascular architecture and hemoglobin oxygenation.

**Methods:** Breast PA imaging was performed in patients with primary breast cancer using a bed-type PA imaging system equipped with a hemispherical sensor and a flat specimen holder enabling mild breast compression. Three independent evaluators assessed predefined characteristics of tumor-associated vasculature: centripetal/disrupted vessels and intratumoral vessel-like signals. Oxygenation (S-factor) of tumor-associated vessels was estimated using dual-wavelength laser irradiation at 756 and 797 nm.

**Results:** PA imaging was performed in 9 tumors from 8 patients. Eight tumors were evaluable, after the exclusion of 1 tumor with segmental bloody discharge. Centripetal/disrupted vessels were identified in 7 tumors (87.5%). Intratumoral vessel-like signals were observed in all tumors (100%), with higher signal density than in surrounding tissue in 5 lesions (62.5%). Increased intratumoral signal density was associated with a higher Ki67-labeling index (two-sided *P* = .01). Mean intratumoral S-factor level (76.9% ± 9.1%) was significantly lower than that of peritumoral vessels at 5 mm (86.4% ± 5.9%) and 20 mm (88.5% ± 4.9%) from the tumor margin (two-sided *P* < .01).

**Conclusion:** PA imaging with a flat specimen holder enables noninvasive visualization of tumor-associated vasculature with reduced oxygenation in primary breast cancer. This approach may provide a novel imaging platform for the early detection and functional assessment of breast cancer.

## Introduction

Abnormal angiogenesis can cause tumor hypoxia, which is a widely recognized hallmark of solid cancers ^1,2^. Because abnormal angiogenesis and tumor hypoxia are associated with aggressive behaviors of solid tumors^3–9^, there is growing demand for the noninvasive visualization of tumor vasculature and its oxygenation. Photoacoustic (PA) imaging is a novel technique that visualizes vessels noninvasively by utilizing the light-absorption properties of hemoglobin ^10–12^. Near-infrared laser light absorbed by hemoglobin induces thermoelastic expansion, which causes PA waves. PA imaging uses the positional information of waves emitted from hemoglobin to visualize the vascular distribution ^13^. Since oxy– and deoxy-hemoglobin have different optical absorption spectra, PA imaging can estimate the oxygen saturation of vessels by using light of different wavelengths (756 and 797 nm) ^14,15^. However, few studies have evaluated the oxygen saturation of primary breast cancer with clear visualization of tumor-associated vasculature in human patients.

The development of PA imaging devices for breast cancer has focused mainly on bed-type and handheld devices ^16,17^. The greatest challenge of PA imaging is that deep blood vessels far from the body surface are difficult to visualize, primarily because near-infrared light is rapidly attenuated in tissues. The handheld type reduces tissue thickness through manual compression. However, manual operation inevitably results in unstable PA signals, and the narrow probe width limits the area in which PA waves can be detected (the “limited view problem”) ^18^. Because it can only obtain 2-dimensional images, evaluations tend to be subjective. Therefore, our group has consistently worked on developing bed-type PA imaging devices.

Our first PA device (PAI-01/02) compressed the breast with 2 plates to allow near-infrared light to reach its deeper part. PAI-01/02 was able to capture tumor-associated PA signals in only 70% of invasive breast cancer cases, and most were visualized as tiny in-tumor spots without detailed vascular structures ^19–21^. Our next models (PAI-03/04) incorporated a hemispherical detector array (HDA), which reduced the limited view problem and enabled precise 3D reconstruction. This array greatly improved visualization performance for body surface vessels, while the breast-holding cup ameliorated patients’ discomfort during the measurement. Despite these advantages, the visualization of the fine structure of blood vessels located in the deeper part of the breasts became more difficult due to the geometric configuration of these models ^11^.

To overcome the drawbacks of these devices, particularly the inability to visualize deeper vessels, we developed LUB-0, a PA device with a flat specimen holder. Mild breast compression from the patient’s own weight enables the visualization of deeper lesions and can expand the imaging range. This model can estimate the tissue hemoglobin oxygen saturation (S-factor) by alternating laser irradiation at 756 and 797 nm. Its excellent performance in visualizing superficial vessels with S-factor information has already been demonstrated^22^. Here, we conducted the first clinical study to evaluate the performance of LUB-0 in characterizing tumor-associated vasculature in human breast cancers. The expanded imaging field, improvements in image processing, and several practical refinements enabled rapid and detailed visualization of tumor-associated vasculature. These advances led to the successful detection of reduced oxygenation within breast tumors through the vascular-level assessment of hemoglobin oxygen saturation, supporting the potential clinical utility of LUB-0.

## Methods

### Ethical guidelines

The study protocol was approved by the Kyoto University Hospital ethics committee (approval number: C1645). Because LUB-0 is structurally identical to LME-01, which has been approved by Japan’s Pharmaceuticals and Medical Devices Agency for use on the body surface (approval number: 30400BZX00212000), its application for breast imaging was considered appropriate. The study period was 4 months due to early termination of the equipment-use agreement associated with the funding period.

### Study participants

The key inclusion criteria were as follows: 1) women with histologically confirmed breast cancer; 2) women aged ≥ 18 years at enrollment; 3) expected ability to tolerate PA imaging without physical discomfort; and 4) written informed consent. The key exclusion criteria were: 1) pregnancy or lactation; 2) regular intake of oral photosensitizing agents for photodynamic therapy (e.g., Photofrin); 3) presence of a cardiac pacemaker; 4) American Society of Anesthesiologists Physical Status Classification class ≥ 4; 5) difficulty maintaining a prone position during PA imaging; and 6) imaging-site infection.

### PA imaging by LUB-0

LUB-0 consists of a patient-imaging bed and a PC console for system control (Fig. 1A). The flat specimen tray (383 [L] × 273 [W] × 16 [D] mm) is placed at the center of the bed, and a HDA is installed beneath (Fig. 1B, C). The pulsed-laser irradiation port is located at the bottom of the HDA. The maximum imaging area is 180 × 290 mm.

**Figure 1.**
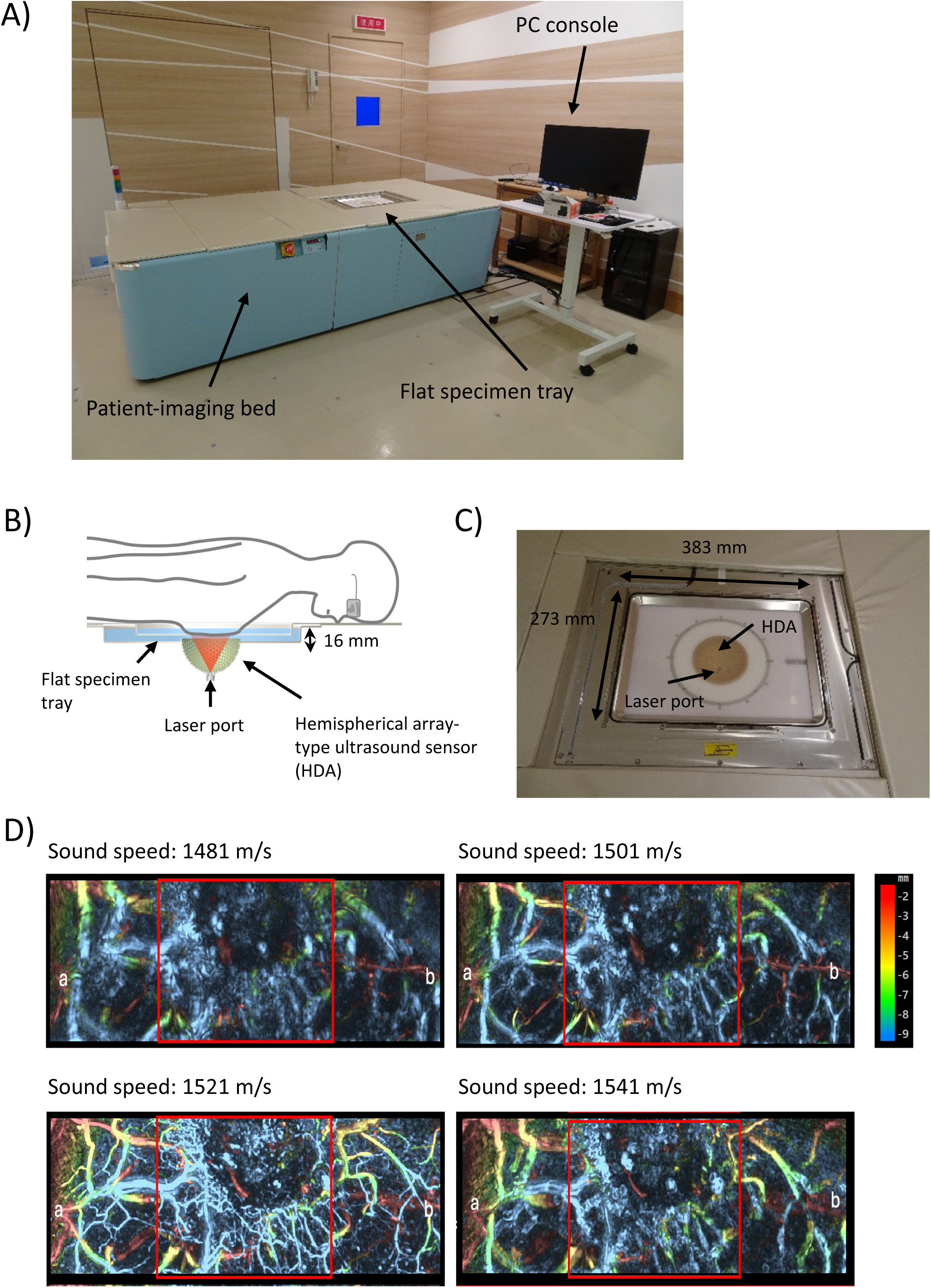
(A) Overview of the LUB-0 photoacoustic (PA) imaging device. A flat specimen tray is located at the center of the patient-imaging bed. (B) Structure of the LUB-0 imaging port. A hemispherical detector array (HDA) is beneath the flat specimen tray. A pulsed-laser port is equipped at the bottom of the HDA. (C) Magnified view of a flat specimen holder. (D) Representative reconstructed PA images (tumor area of patient 1) created by using different sound speed settings. Sound speed for calculation was set at 1481 m/s (upper right), 1501 m/s (upper left), 1521 m/s (lower right), and 1541 m/s (lower left). **Alt text:** Pictures showing the configuration of the LUB-0 photoacoustic imaging device and representative reconstructed images. The LUB-0 has a flat specimen tray at the center of the patient-imaging bed, where the breast is irradiated with pulsed-laser from the bottom of a hemispherical detector array. The user can adjust assumed sound speed to obtain the highest-resolution photoacoustic image.

Before PA imaging, the tumor location was marked on the skin with a purple pen (P-MO-120-MC-PU oil-based marker Mackee; Zebra, Tokyo, Japan) via a breast ultrasound examination (HiVISION Ascendus; Hitachi Medico, Tokyo, Japan) with the patient in the supine position. Then, the patient lay in the prone position and raised 1 arm to insert 1 breast into the rectangular specimen holder. The HDA and specimen holder were filled with water at a temperature of approximately 30°C (Fig. 1C). Pulsed laser with different wavelengths (756 and 797 nm) was alternately irradiated to the breast at 0.033-s intervals (30 Hz), and the emitted PA signals were captured by the HDA. Volume data were acquired at multiple angular positions during scanning and reconstructed into a 3D image. After whole-breast scanning in standard mode, tumor-area scanning in high-quality mode was performed in the affected breast.

Universal Back Projection was used for 3D image reconstruction^23^, with a reconstructed volumetric-data voxel size of 0.1 mm. In the 3D reconstruction process, we adjusted the assumed sound speeds in 5 m/s increments around the default value and selected the value yielding the sharpest and finest vascular structures in the region of interest (Fig. 1D). Motion artifacts during imaging were corrected as previously reported^11^. A dedicated viewer (PAT Viewer, Luxonus Inc., Kawagawa, Japan) was used for assessing PA images.

### Evaluation of deep-vessel imaging performance

To evaluate the performance of LUB-0 for imaging deep vessels, we measured the distance from the skin surface to the deepest vessels visualized by PA imaging. This distance was compared with the estimated breast thickness, defined as the distance from the skin surface to the edge of the pectoral muscle on breast ultrasonography.

### Evaluation of the visualization of tumor-associated vasculature

Two qualified breast surgeons and 1 qualified radiologist evaluated the performance of LUB-01 for visualizing tumor-associated vasculature. Based on our previous studies, the evaluators assessed the presence or absence of the following 3 features that characterize tumor-associated vasculature in PA images: 1) centripetally directed toward the tumor, 2) disruption at the tumor boundary, and 3) intratumoral signals with linear/spotty structure. Each evaluator scored the 3 features as “Definitely present (score = 2)”, “Possibly present but uncertain (score = 1)”, and “Not present (score = 0)”. Total scores of the 3 evaluators ≥ 5 for each item were rated “present.” In addition to these key features, the PA signal density inside the tumors was compared with that of surrounding normal areas and scored as higher (score = 2), equal (score = 1), or lower (score = 0). Total scores of 0–3 and 4–6 were interpreted as low and high density, respectively. We evaluated the associations of the PA signal density with tumor pathological factors (tumor size, histological grade, estrogen receptor [ER], progesterone receptor [PgR], HER2, Ki67) and patient backgrounds (age, body mass index [BMI], tumor depth from skin).

### S-factor measurement

To evaluate oxygenation levels, the S-factor of tumor-associated vessels was calculated from the data acquired using alternate-laser irradiation with 2 different wavelengths, as previously described ^21,24,25^. The S-factors for intratumoral vessels and for peritumoral vessels (5 mm and 20 mm from the tumor margin) were compared at the same depth. The S-factors of the intratumoral vessels were acquired at 2-mm intervals. S-factors were extracted at least 3 times from each measurement point to minimize errors due to manual point placement.

### Statistical analysis and software

Chi-square tests were used for the analysis of categorical variables, specifically the associations between PA signal density and clinicopathological factors. Mann-Whitney tests with Benjamini, Krieger, and Yektieli correction for multiple comparisons were used to evaluate differences in S-factors. All statistical tests were two-sided, and a *P* value < .05 was considered statistically significant. GraphPad Prism ver. 10 was used for all statistical analyses and to plot the data. The illustration of Fig. 3A was created using BioRender application. The manuscript was edited by a professional language editing service and reviewed by all authors.

## Results

### Participant and breast tumor characteristics

Eight women with breast cancer were enrolled during the study period. The participants’ background characteristics and the histopathological characteristics of their tumors are presented in Table 1. The median age of the patients was 64.5 (range, 54–85) years, and their median BMI was 21 (range, 18–25). Because 1 participant (Patient 7) had 2 primary lesions in the same breast, a total of 9 tumors were evaluated. All tumors were invasive breast cancer: 8 were hormone receptor (HR)-positive/HER2-negative, and 1 was HR-positive/HER2-positive. Except for Patient 8, who had renal dysfunction, all patients underwent contrast-enhanced breast MRI. Skin redness without ulceration was observed in Patient 1. Bloody discharge in the mammary ducts was noted in the segment occupied by the tumor in Patient 3. All tumors had irregular margins on breast ultrasonography, and at least 1 flow signal was confirmed on Doppler ultrasound, except in Patient 2.

**Table 1.**
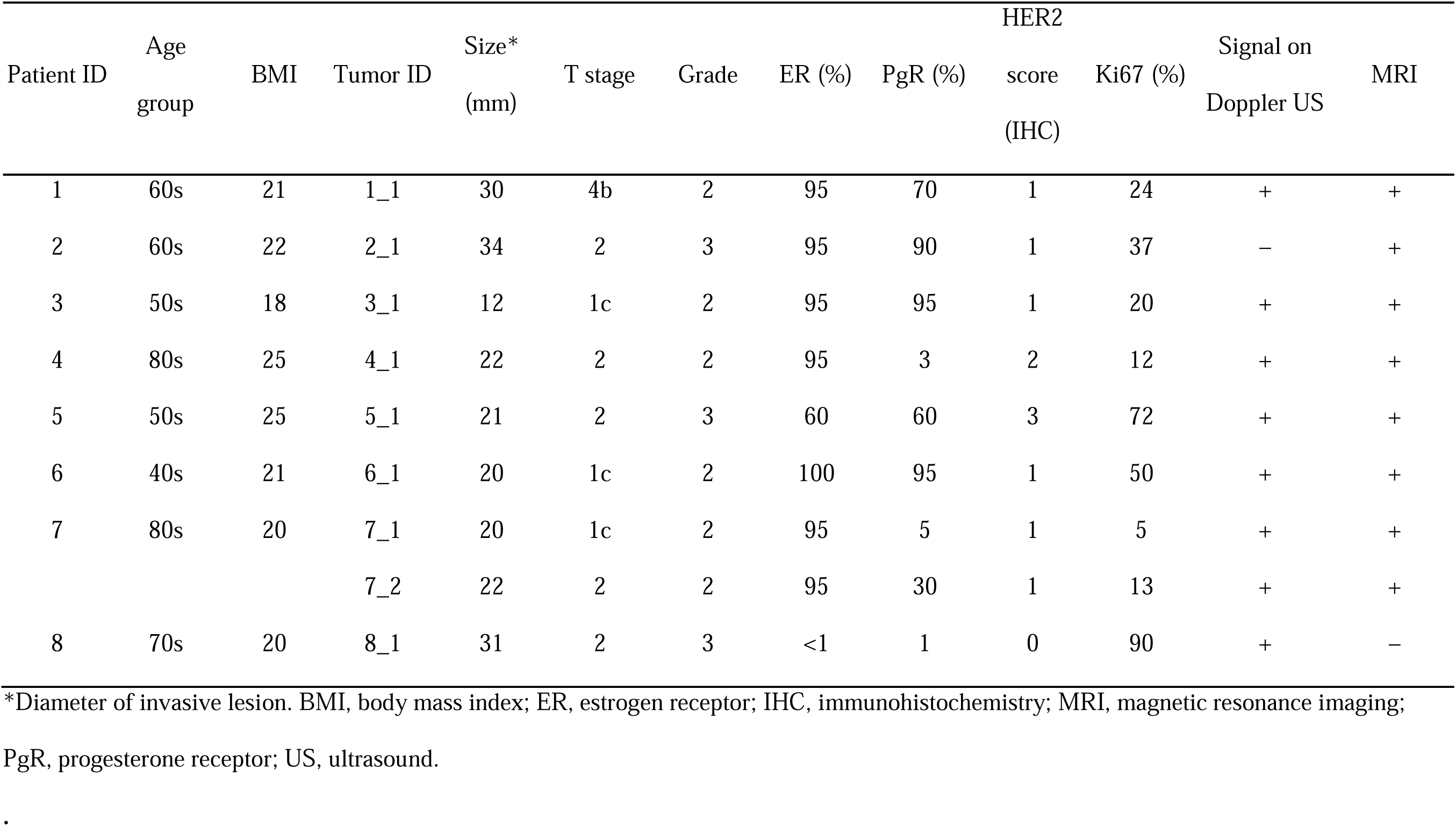
Key characteristics of participants and tumors.

| Patient ID | Age group | BMI | Tumor ID | Size* (mm) | T stage | Grade | ER (%) | PgR (%) | HER2 score (IHC) | Ki67 (%) | Signal on Doppler US | MRI |
| --- | --- | --- | --- | --- | --- | --- | --- | --- | --- | --- | --- | --- |
| 1 | 60s | 21 | 1_1 | 30 | 4b | 2 | 95 | 70 | 1 | 24 | + | + |
| 2 | 60s | 22 | 2_1 | 34 | 2 | 3 | 95 | 90 | 1 | 37 | – | + |
| 3 | 50s | 18 | 3_1 | 12 | 1c | 2 | 95 | 95 | 1 | 20 | + | + |
| 4 | 80s | 25 | 4_1 | 22 | 2 | 2 | 95 | 3 | 2 | 12 | + | + |
| 5 | 50s | 25 | 5_1 | 21 | 2 | 3 | 60 | 60 | 3 | 72 | + | + |
| 6 | 40s | 21 | 6_1 | 20 | 1c | 2 | 100 | 95 | 1 | 50 | + | + |
| 7 | 80s | 20 | 7_1 | 20 | 1c | 2 | 95 | 5 | 1 | 5 | + | + |
|  |  |  | 7_2 | 22 | 2 | 2 | 95 | 30 | 1 | 13 | + | + |
| 8 | 70s | 20 | 8_1 | 31 | 2 | 3 | <1 | 1 | 0 | 90 | + | – |

\*Diameter of invasive lesion. BMI, body mass index; ER, estrogen receptor; IHC, immunohistochemistry; MRI, magnetic resonance imaging;
PgR, progesterone receptor; US, ultrasound.

### Detection limit and tolerability of PA imaging

We evaluated the performance of LUB-0 for visualizing deep-tissue vessels. Table 2 shows the deepest positions of blood vessels in PA imaging and their ratios to the estimated breast thickness on ultrasonography. On average, vessels ≤12.9-mm deep could be detected by PA imaging (highest in Patient 1: 17.6 ± 3.5 mm in 1 case). The average ratio to breast thickness was 84% (highest in Patinet 7: 111% ± 6%). In the breast-region comparisons, the range of PA imaging depth tended to be greater in the upper-outer quadrant (average depth, 14 ± 2.9 mm), while the coverage ratio tended to be higher in the lower-outer quadrant.

**Table 2.** Maximum detection depth of photoacoustic signals.

| Patient ID | Visible depth |  | Ratio to breast thickness |  | Breast Area | Visible depth |  | Ratio to breast thickness |  |
| --- | --- | --- | --- | --- | --- | --- | --- | --- | --- |
|  | Depth (mm) | SD | (%) | SD |  | Depth (mm) | SD | (%) | SD |
| 1 | 17.6 | ±3.5 | 81% | ±20 | Upper inner | 13.5 | ±2.5 | 84 | ±14 |
| 2 | 12.6 | ±1.7 | 78% | ±12 | Upper outer | 14 | ±2.9 | 76 | ±18 |
| 3 | 10.5 | ±1.6 | 92% | ±8 | Lower inner | 11.1 | ±1.2 | 82 | ±22 |
| 4 | 12.5 | ±3.4 | 67% | ±17 | Lower outer | 12.9 | ±2.4 | 95 | ±11 |
| 5 | 14.0 | ±1.9 | 76% | ±8 |  |  |  |  |  |
| 6 | 11.6 | ±1.7 | 78% | ±18 |  |  |  |  |  |
| 7 | 11.3 | ±1.0 | 111% | ±6 |  |  |  |  |  |
| 8 | 13.1 | ±1.2 | 93% | ±6 |  |  |  |  |  |
| Average | 12.9 |  | 84% |  |  |  |  |  |  |

SD, standard deviation.

The average imaging time for an entire breast was 2-3 minutes in standard mode and 5-7 minutes for the tumor area in high-resolution mode. No participants complained of discomfort associated with the water temperature or maintaining their posture, and no adverse events were observed related to the PA imaging. PA imaging was successful in both younger and older patients (Patients 4 and 7: >80 years old).

### Detection of tumor-associated vasculature

The patient with abundant bloody discharge (Patient 3) was excluded from the evaluation because the PA signals from the discharge masked the tumor-associated vasculature. Consequently, tumor-associated vasculature was evaluated in 8 tumors from 7 patients.

The mean score for “centripetal” vessels toward the tumor was 5.4; they were judged as “present” in 7 of the 8 lesions (87.5%) (Table 3). The mean score for “disruption” at the tumor margin was 5.0; they were judged as “present” in 7 of the 8 lesions (87.5%). The mean score for “intratumoral signals” with linear/spotty structure was 5.25; they were judged as “present” in all lesions (100%). Representative PA images are shown in which all evaluators agreed that all 3 features were present (Fig. 2A, B; Tumors 1-1 and 5-1). All tumors exhibited at least 2 of the 3 evaluated features, indicating excellent performance in detecting tumor-associated vessels.

**Figure 2.**
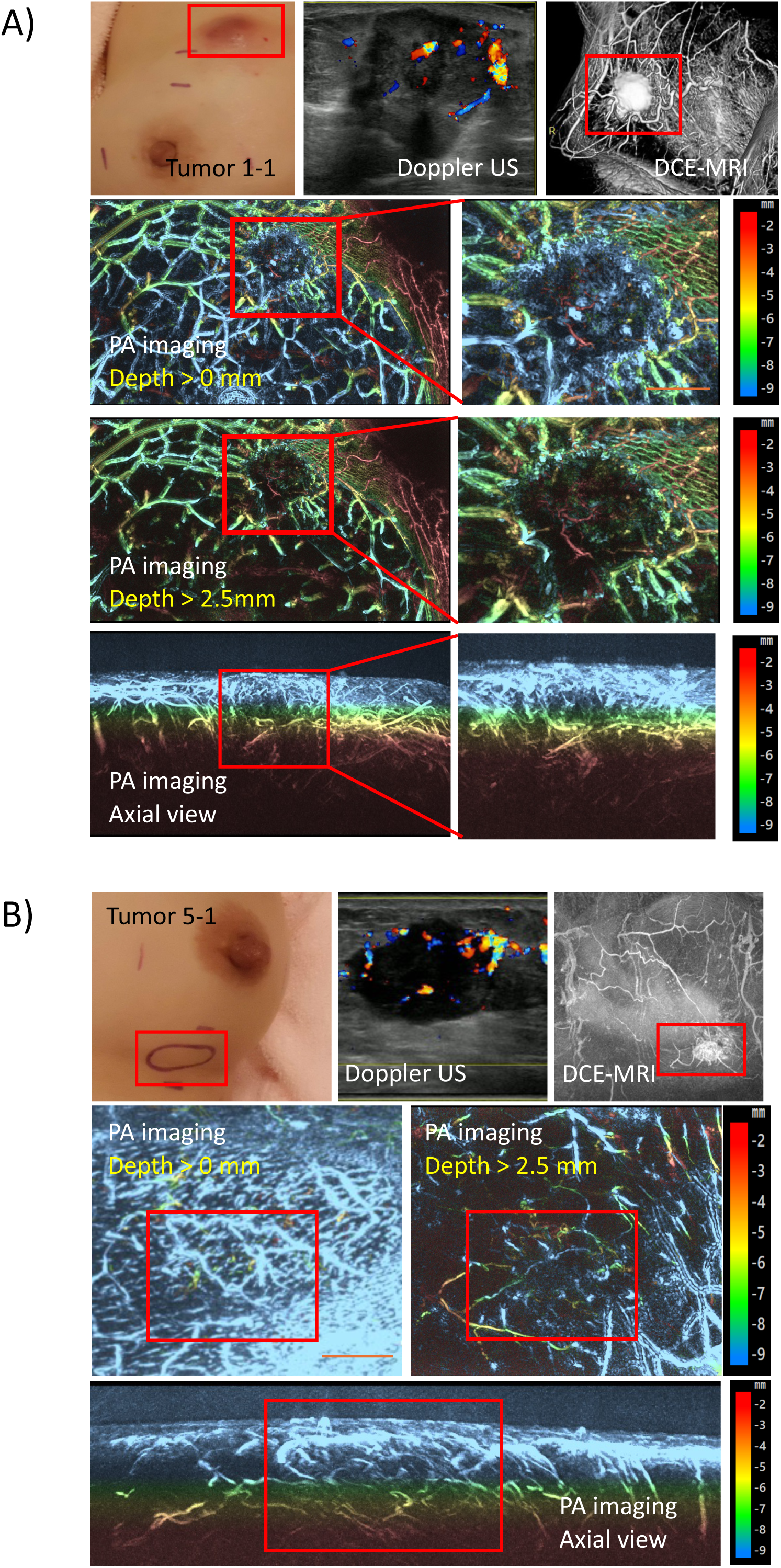
Representative photoacoustic (PA) images of (A) Tumor 1–1 (Patient 1) and (B) Tumor 5–1 (Patient 5). The upper panels show a clinical photograph of the breast lesion, a Doppler ultrasound image, and a dynamic contrast-enhanced (DCE) MRI image. The middle panels show images reconstructed using signals from depths greater than 2.5 mm. The lower panels show PA images of the affected breast. The color key indicates vessel depth from the skin surface. Scale bar, 10 mm. **Alt text:** Composite figures showing clinical photographs of the affected breast, Doppler ultrasound images, dynamic contrast-enhanced MRI image and corresponding photoacoustic images of Tumors 1-1 and 5-1. In both tumors, vessels running toward the center of the tumor are clearly visualized in images reconstructed using the signals from depth greater than 2.5 mm. The centripetal vessels are disrupted at the tumor boundary, and discontinuous linear vessel-like– and dot-like signals are observed within the tumors.

**Figure 3.**
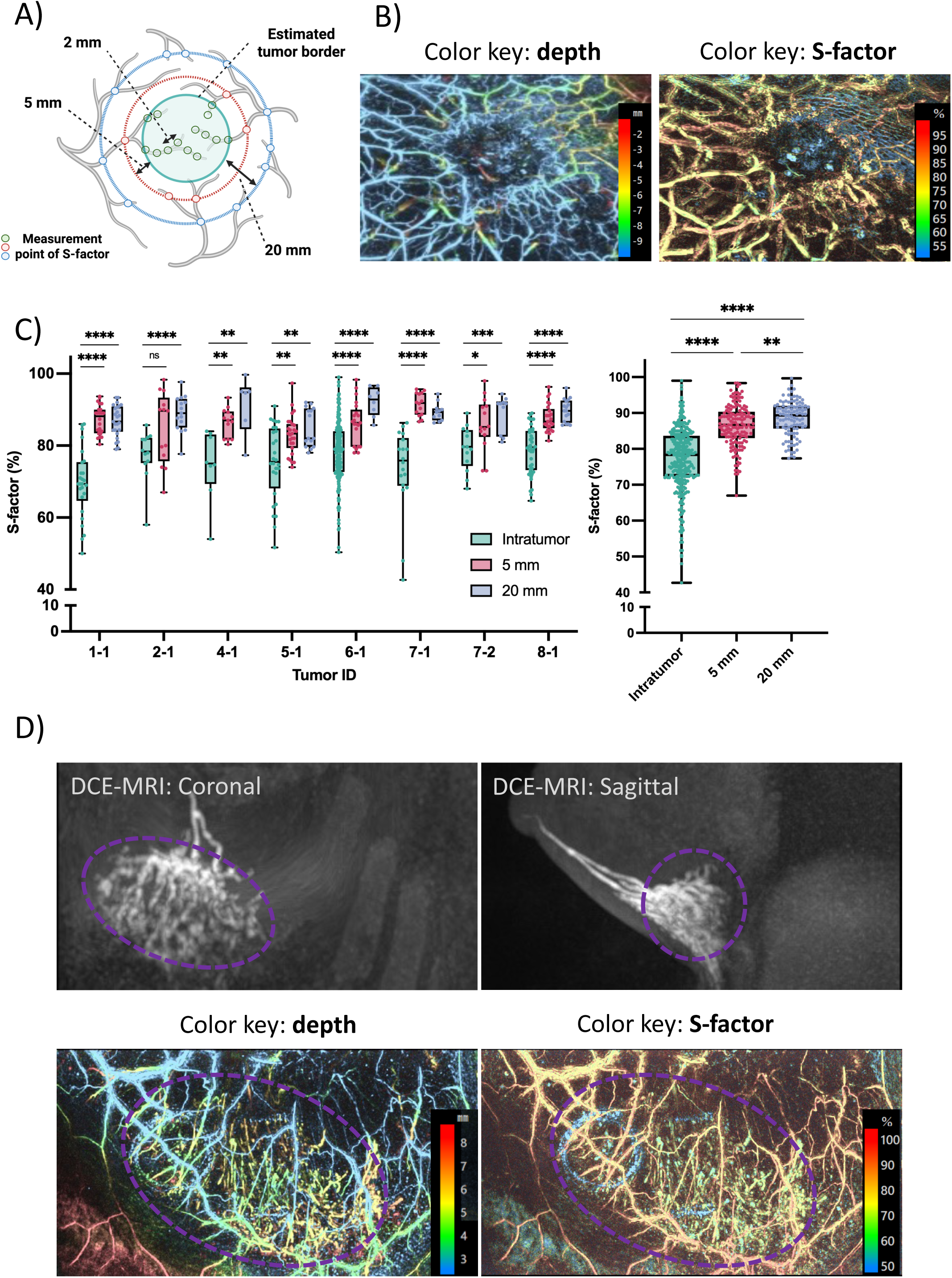
(A) S-factors within the tumor (inside the green circle) and at 5 mm (red circle) and 20 mm (blue circle) from the tumor border were evaluated. (B) Representative photoacoustic (PA) image of tumor-associated vasculature (left) and the corresponding S-factor projection (right). The color keys in the left and right panels indicate vessel depth and estimated S-factor, respectively. (C) S-factor differences at each measurement point. Left: S-factors of tumor-associated vasculature in evaluable tumors. Right: summary data of S-factors within tumors and at 5 mm and 20 mm from the tumor borders. Box plots show the median and 25th and 75th percentiles; whiskers indicate the minimum and maximum values. \**P* < .05, \*\**P* < .01, \*\*\**P* < .001, \*\*\*\**P* < .0001; n.s., not significant. (D) Upper panels show dynamic contrast-enhanced MRI (DCE-MRI) images of Tumor 3_1 (Patient 3). The dotted lines indicate segmental accumulation of bloody discharge. Lower panels show the corresponding PA image (left) and S-factor projection (right). Diffuse linear PA signals with lower S-factors were observed in the area of bloody discharge. **Alt text**: Figures and graphs showing reduced S-factors within tumors and in an area of bloody discharge. The intratumoral S-factor levels are significantly lower than those of peritumoral vessels at 5 mm and 20 mm from the tumor border in all evaluable tumors. Reduction of S-factor associated with segmental bloody discharge is also visualized.

**Table 3.**
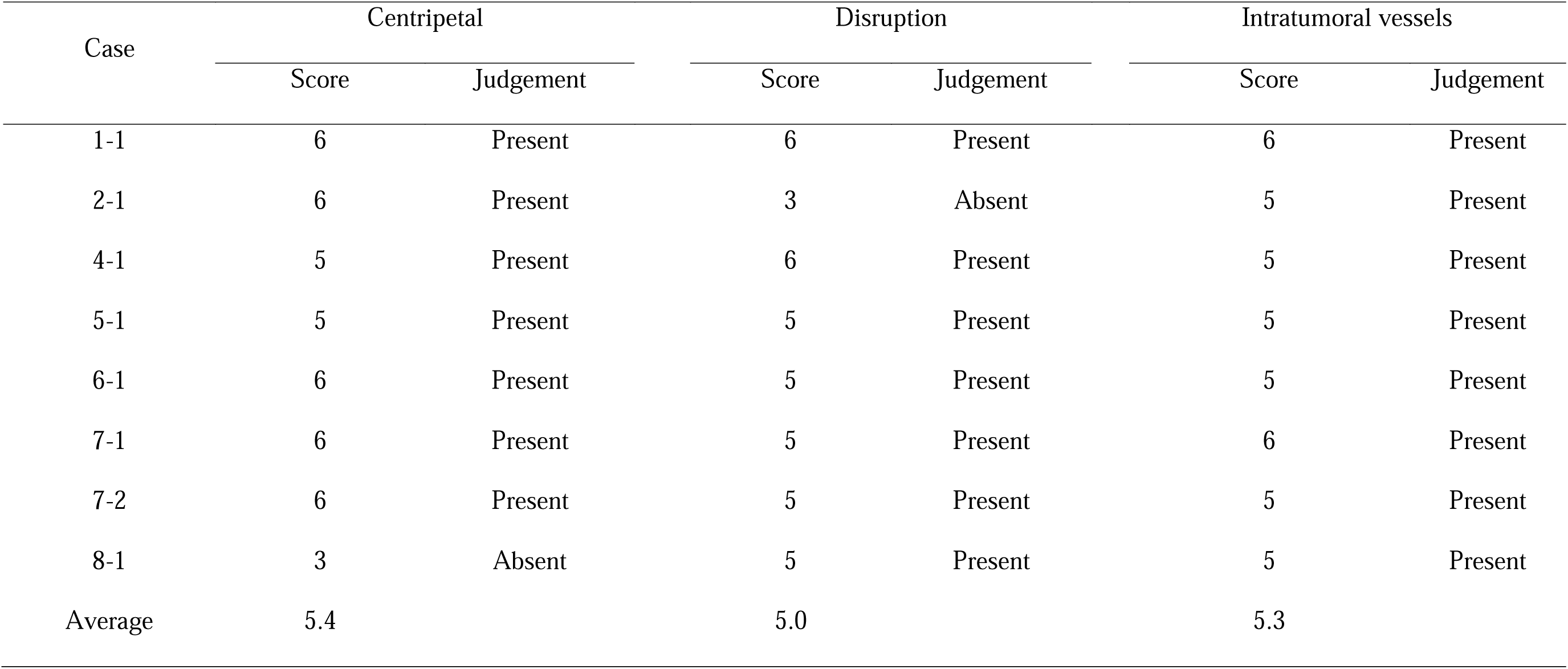
Evaluation of key features related to tumor-associated vasculature.

In the comparison between tumors and surrounding normal tissues, the PA signal density was assessed as “higher in the tumor” in 5 lesions (62.5%) and as “lower in the tumor” in 3 (37.5%). The 2 lesions judged as lower were from the same patient. Higher tumor PA signal density was significantly associated with a higher Ki67 labeling index (Table 4). This finding reflected the fact that highly proliferative tumors exhibit a more angiogenic signature.

**Table 4.** Association between PA signal density and tumor characteristics.

| Characteristic |  | Intratumoral PA signal density |  | P value |
| --- | --- | --- | --- | --- |
|  |  | High (n = 5) | Low (n = 3) |  |
| Tumor size (mm) | ≤20 | 1 | 2 | .46 |
|  | >20 | 4 | 1 |  |
| Grade | 1, 2 | 2 | 3 | .19 |
|  | 3 | 3 | 0 |  |
| ER* | + | 4 | 3 | >.99 |
|  | - | 1 | 0 |  |
| PgR* | + | 4 | 1 | .46 |
|  | - | 1 | 2 |  |
| HER2 | + | 1 | 0 | >.99 |
|  | - | 4 | 3 |  |
| Ki67** | High | 5 | 0 | .01 |
|  | Low | 0 | 3 |  |
| Signal on Doppler | + | 4 | 3 | >.99 |
| US | - | 1 | 0 |  |

\*Threshold for positivity is 10% or over. \*\*Threshold for positivity is 15% or over. ER, estrogen receptor; PA, photoacoustic; PgR, progesterone receptor; US, ultrasound.

### Oxygen saturation of tumor-associated vasculature

Finally, we compared S-factor values inside the tumor and at 5 mm and 20 mm from the tumor border to evaluate the oxygen saturation in tumor-associated vasculature (Fig. 3A). In all 8 evaluable tumors, the S-factor was significantly lower inside the tumor (mean intratumoral S-factor: 76.9% ± 9.1%) than in the peritumoral vessels at 20 mm from the tumor border (mean peritumoral S-factor at 20 mm: 88.5% ± 4.9%) (Fig. 3B, C). The S-factor was also lower inside the tumor than at 5 mm from the tumor border in 7 out of 8 tumors (mean peritumoral S-factor at 5mm: 86.4% ± 5.9%). The S-factor at 5 mm from tumor border was numerically lower than that at 20 mm although the differences were not statistically significant. The S-factor variation tended to be greater inside tumors than in the peritumoral vessels, implicating an effect of the abnormal vessels within the tumors on oxygen saturation (Fig. 3C).

All images of tumor-associated vessels and their S-factor are displayed in Fig. S1.

As an exploratory analysis, we evaluated the S-factor in the bloody discharge of Patient 3. The average S-factor in the bloody discharge was 59.8% compared with 86.8% in the vascular portion of the same segment (Fig. 3D). This finding is consistent with the expected lower oxygenation of blood within the discharge, supporting the validity of the S-factor as a measure of hemoglobin oxygenation.

## Discussion

In the present study of 9 women with breast cancer, the bed-type PA imaging device, LUB-0, successfully visualized the characteristic patterns of the tumor-associated vasculature in all evaluated tumors with improved resolution compared with our previous models. Importantly, we observed a significant and consistent reduction in S-factor, a measure of oxygenation, inside the tumors. This is the first demonstration of reduced oxygenation in human breast tumors based on S-factor assessment on clearly visualized tumor-associated vasculature using bed-type PA imaging. These findings highlight the advances of LUB-0 over to the earlier PA imaging systems and support its clinical application as a novel functional imaging modality for breast tumors.

Multiple studies using near-infrared diffuse optical tomography (NIR-DOT) showed that the assessment of oxygenation levels in primary breast tissue could be useful for predicting tumor behavior, such as proliferative capacity and chemotherapy sensitivity ^26–29^. The positive findings with NIR-DOT prompted efforts to develop devices for the clinical assessment of tumor oxygenation. PA imaging devices can localize tumors more precisely than NIR-DOT, enabling targeted assessment of tumor oxygenation status. Indeed, some clinical studies using handheld devices reported reduced oxygenation in cancer tissue compared with benign tissue^30–35^. Our successful detection of an S-factor reduction at intratumoral vessels using LUB-0 further supports the clinical implementation of PA imaging devices to acquire additional information that can predict tumor aggressiveness. A comparison of the clinicopathological characteristics of breast tumors revealed a positive correlation between the density of intratumoral PA signals and the Ki67 value, consistent with our previous study, in which we reported a correlation between increased vascular branching and higher Ki67 values^36^. These findings support the value of PA imaging in predicting the proliferation capacity of tumors.

In addition to the functional assessment of primary tumors, PA imaging is expected to be applied for noninvasive breast screening ^7^. We previously defined 3 representative features of tumor-associated vessels observed via PA imaging of breast cancer: 1) centripetally directed toward the tumor, 2) disruption of vasculature at the tumor margin, and 3) fine spotty or linear intratumoral PA signals. In the trials evaluating 22 invasive breast cancers with a previous model (PAI-03), “centripetal” and/or “disruption” was confirmed in less than 70% of the cases, and PA signals within the tumor were detected in only half of the cases^11^. Intratumoral oxygenation was successfully identified in limited cases^25^. In the present study, at least 2 of these features were observed in all lesions, and intratumoral PA signals showed distinct vessel-like structure in several cases. Moreover, S-factor reductions were observed at intratumoral regions in all cases. Although all evaluators agreed that it was difficult to identify the tumor solely based on the vessel structures, even with LUB-0, its improved performance in capturing tumor-associated vessels and their S-factors may contribute to the development of a robust breast cancer-screening system by combining structural information of the vessels with oxygenation status.

A key reason for this improvement is the use of tissue compression ^37^. The shallow and flat specimen holder leads to mild breast compression under the patient’s own weight, which can minimize the attenuation of near-infrared light by reducing tissue thickness, thereby improving detectability. Recently, a PA imaging device with similar geometry showed favorable diagnostic ability for primary breast cancer^38^. However, because this device uses a single laser with a longer wavelength (1064 nm) to achieve deeper tissue penetration, oxygen saturation cannot be estimated. Another reason for the improved performance of this model is the incorporation of several image correction procedures to reduce motion artifacts. Importantly, selecting the optimal sound speed for image reconstruction also enabled higher resolution imaging because the sound speed depends on the lipid composition of the tissue.

This study has several limitations. First, the number of cases included was small because of the limited study period. The small sample size particularly made it difficult to examine the association between PA signals and important clinical factors, such as associations with ER, HER2, and BMI, with sufficient statistical power. Second, the study subjects were all Japanese women evaluated at a single institution. Therefore, validation studies including multiple institutions and women from diverse racial and ethnic backgrounds are warranted. Third, we did not verify the findings regarding intratumoral PA signals and their oxygenation by other methods. Although the accuracy of S-factor measured by LUB-0 has previously been confirmed in tumor-bearing mouse models^39^), it should be noted that the S-factor used in the study does not represent the absolute quantification of tissue oxygenation.

In conclusion, our new PA device equipped with a flat specimen holder substantially improves the visualization of tumor-associated vessels in breast cancer. The notable improvement in the recognition of intratumoral PA signals, together with the significant reduction in estimated oxygen saturation, adds to the growing body of evidence supporting the clinical application of PA imaging, particularly for the functional assessment of breast cancer and noninvasive cancer screening.

## Acknowledgements

The sponsor did not play a role in the design of the study; the collection, analysis, and interpretation of the data; the writing of the manuscript; and the decision to submit the manuscript for publication.

## Data availability

The primary data of human subjects cannot be provided without appropriate data transfer agreement and additional consents from participants. Analyzed data are available from the corresponding author upon reasonable request.

## References

1. Hanahan D, Coussens LM. Accessories to the crime: functions of cells recruited to the tumor microenvironment. Cancer Cell. 2012;21(3):309–322.

2. Hanahan D, Weinberg RA. The Hallmarks of Cancer Review evolve progressively from normalcy via a series of pre.

3. Harris AL. Hypoxia--a key regulatory factor in tumour growth. Nat Rev Cancer. 2002;2(1):38–47.

4. Choi WWL, Lewis MM, Lawson D, et al. Angiogenic and lymphangiogenic microvessel density in breast carcinoma: correlation with clinicopathologic parameters and VEGF-family gene expression. Mod Pathol. 2005;18(1):143–152.

5. Hansen S, Grabau DA, Sørensen FB, Bak M, Vach W, Rose C. The prognostic value of angiogenesis by Chalkley counting in a confirmatory study design on 836 breast cancer patients. Clin Cancer Res. 2000;6(1):139–146.

6. Bosari S, Lee AK, DeLellis RA, Wiley BD, Heatley GJ, Silverman ML. Microvessel quantitation and prognosis in invasive breast carcinoma. Hum Pathol. 1992;23(7):755–761.

7. Nagy JA, Chang SH, Dvorak AM, Dvorak HF. Why are tumour blood vessels abnormal and why is it important to know? Br J Cancer. 2009;100(6):865–869.

8. Toi M, Inada K, Suzuki H, Tominaga T. Tumor angiogenesis in breast cancer: its importance as a prognostic indicator and the association with vascular endothelial growth factor expression. Breast Cancer Res Treat. 1995;36(2):193–204.

9. Tsutsui S, Kume M, Era S. Prognostic value of microvessel density in invasive ductal carcinoma of the breast. Breast Cancer. 2003;10(4):312–319.

10. Quarto G, Spinelli L, Pifferi A, et al. Estimate of tissue composition in malignant and benign breast lesions by time-domain optical mammography. Biomed Opt Express. 2014;5(10):3684–3698.

11. Toi M, Asao Y, Matsumoto Y, et al. Visualization of tumor-related blood vessels in human breast by photoacoustic imaging system with a hemispherical detector array. Sci Rep. 2017;7(1):41970.

12. Wang LV, Hu S. Photoacoustic tomography: in vivo imaging from organelles to organs. Science. 2012;335(6075):1458–1462.

13. Wang LV, Yao J. A practical guide to photoacoustic tomography in the life sciences. Nat Methods. 2016;13(8):627–638.

14. Wang X, Xie X, Ku G, Wang LV, Stoica G. Noninvasive imaging of hemoglobin concentration and oxygenation in the rat brain using high-resolution photoacoustic tomography. J Biomed Opt. 2006;11(2):024015.

15. Asao Y, Hirano R, Nagae K, et al. Visualization of spatial distribution of hemoglobin with various oxygen saturations in small animals using a photoacoustic imaging scanner with a hemispherical detector array. bioRxiv. Published online 2023. doi:10.1101/2023.06.19.545650

16. Manohar S, Dantuma M. Current and future trends in photoacoustic breast imaging. Photoacoustics. 2019;16(100134):100134.

17. Nyayapathi N, Xia J. Photoacoustic imaging of breast cancer: a mini review of system design and image features. J Biomed Opt. 2019;24(12):1–13.

18. Xu Y, Wang LV, Ambartsoumian G, Kuchment P. Reconstructions in limited-view thermoacoustic tomography. Med Phys. 2004;31(4):724–733.

19. Kitai T, Torii M, Sugie T, et al. Photoacoustic mammography: initial clinical results. Breast Cancer. 2014;21(2):146–153.

20. Fakhrejahani E, Torii M, Kitai T, et al. Clinical report on the first prototype of a photoacoustic tomography system with dual illumination for breast cancer imaging. PLoS One. 2015;10(10):e0139113.

21. Asao Y, Hashizume Y, Suita T, et al. Photoacoustic mammography capable of simultaneously acquiring photoacoustic and ultrasound images. J Biomed Opt. 2016;21(11):116009.

22. Tsuge I, Munisso MC, Kosaka T, et al. Preoperative visualization of midline-crossing subcutaneous arteries in transverse abdominal flaps using photoacoustic tomography. J Plast Reconstr Aesthet Surg. 2023;84:165–175.

23. Xu M, Wang LV. Universal back-projection algorithm for photoacoustic computed tomography. Phys Rev E Stat Nonlin Soft Matter Phys. 2005;71(1 Pt 2):016706.

24. Shiina T, Toi M, Yagi T. Development and clinical translation of photoacoustic mammography. Biomed Eng Lett. 2018;8(2):157–165.

25. Matsumoto Y, Asao Y, Sekiguchi H, et al. Visualising peripheral arterioles and venules through high-resolution and large-area photoacoustic imaging. Sci Rep. 2018;8(1):14930.

26. Nioka S, Chance B. NIR spectroscopic detection of breast cancer. Technol Cancer Res Treat. 2005;4(5):497–512.

27. Brown JQ, Wilke LG, Geradts J, Kennedy SA, Palmer GM, Ramanujam N. Quantitative optical spectroscopy: a robust tool for direct measurement of breast cancer vascular oxygenation and total hemoglobin content in vivo. Cancer Res. 2009;69(7):2919–2926.

28. Ueda S, Roblyer D, Cerussi A, et al. Baseline tumor oxygen saturation correlates with a pathologic complete response in breast cancer patients undergoing neoadjuvant chemotherapy. Cancer Res. 2012;72(17):4318–4328.

29. Cochran JM, Busch DR, Leproux A, et al. Tissue oxygen saturation predicts response to breast cancer neoadjuvant chemotherapy within 10 days of treatment. J Biomed Opt. 2018;24(2):1–11.

30. Huang Z, Tian H, Luo H, et al. Assessment of oxygen saturation in breast lesions using Photoacoustic imaging: Correlation with benign and malignant disease. Clin Breast Cancer. 2024;24(4):e210–e218.e1.

31. Nandy S, Mostafa A, Hagemann IS, et al. Evaluation of ovarian cancer: Initial application of coregistered photoacoustic tomography and US. Radiology. 2018;289(3):740–747.

32. Yang M, Zhao L, He X, et al. Photoacoustic/ultrasound dual imaging of human thyroid cancers: an initial clinical study. Biomed Opt Express. 2017;8(7):3449–3457.

33. Kim J, Park B, Ha J, et al. Multiparametric photoacoustic analysis of human thyroid cancers in vivo. Cancer Res. 2021;81(18):4849–4860.

34. Zhang R, Zhao LY, Zhao CY, et al. Exploring the diagnostic value of photoacoustic imaging for breast cancer: the identification of regional photoacoustic signal differences of breast tumors. Biomed Opt Express. 2021;12(3):1407–1421.

35. Neuschler EI, Butler R, Young CA, et al. A pivotal study of optoacoustic imaging to diagnose benign and malignant breast masses: A new evaluation tool for radiologists. Radiology. 2018;287(2):398–412.

36. Yamaga I, Kawaguchi-Sakita N, Asao Y, et al. Vascular branching point counts using photoacoustic imaging in the superficial layer of the breast: A potential biomarker for breast cancer. Photoacoustics. 2018;11:6–13.

37. Kitai T, Kawashima M. Transcutaneous detection and direct approach to the sentinel node using axillary compression technique in ICG fluorescence-navigated sentinel node biopsy for breast cancer. Breast Cancer. 2012;19(4):343–348.

38. Huang K, Fu P, Zhu H, et al. High-speed photoacoustic and ultrasonic computed tomography of the breast tumor for early diagnosis with enhanced accuracy. Sci Adv. 2025;11(41):eadz2046.

39. Kruger RA, Kuzmiak CM, Lam RB, Reinecke DR, Del Rio SP, Steed D. Dedicated 3D photoacoustic breast imaging: Dedicated 3D photoacoustic breast imaging. Med Phys. 2013;40(11):113301.

