## Supplementary material for "Detection of intratumoral hypoxia in primary breast cancer using photoacoustic imaging": Fig. S1

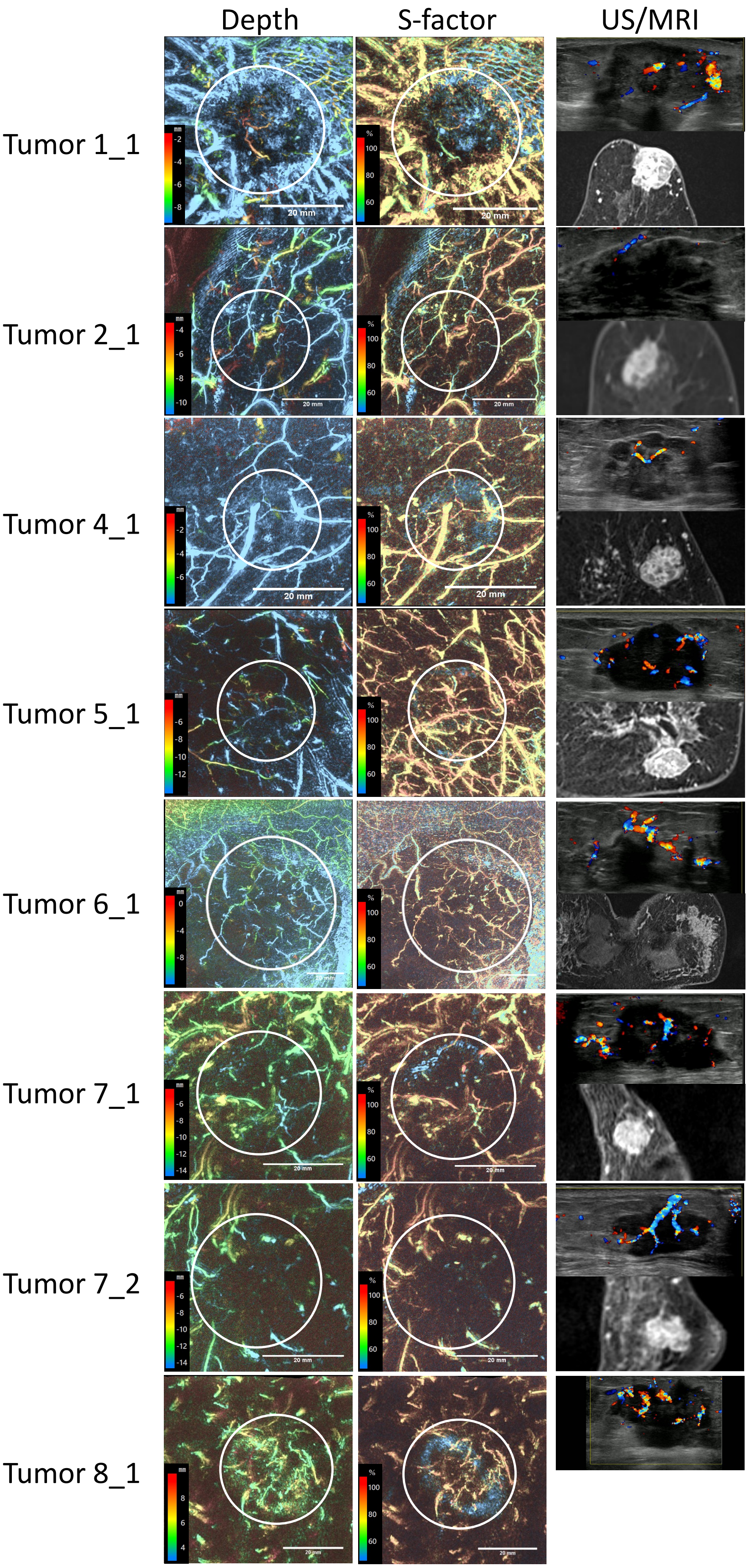


Fig. S1

PA images of all 8 evaluable tumors. The left and middle panels show PA images of tumor-associated vasculature and the corresponding S-factor projection, respectively. The color keys in the left and middle panels indicate vessel depth and the estimated S-factor, respectively. The right panels show Doppler ultrasound images (upper) and dynamic contrast-enhanced MRI images (lower). Round circles indicate the estimated tumor boundary. US, ultrasound.
